# Manufacturer Signal-to-Cutoff Threshold Underestimates Cumulative Incidence of SARS-CoV-2 Infection: Evidence from the Los Angeles Firefighters Study

**DOI:** 10.1101/2021.04.20.21255829

**Authors:** Omar Toubat, Anders H. Berg, Kimia Sobhani, Karen Mulligan, Acacia M. Hori, Jay Bhattacharya, Neeraj Sood

**Author notes:** **Address for Correspondence:** Neeraj Sood, PhD, Professor of Public Policy, Verna and Peter Dauterive Hall, University of Southern California, 635 Downey Way, Los Angeles, CA 90089.

## Abstract

While SARS-CoV-2 serologic testing is used to measure cumulative incidence of COVID-19, appropriate signal-to-cut off (S/Co) thresholds remain unclear. We demonstrate S/Co thresholds based on known negative samples significantly increases seropositivity and more accurately estimates cumulative incidence of disease compared to manufacturer-based thresholds.

## Introduction

Designing an appropriate public health response to the pandemic requires an accurate estimate of the cumulative incidence of severe acute respiratory syndrome coronavirus 2 (SARS-CoV-2) infection. While serologic tests for antibodies against SARS-CoV-2 have served as the primary method for modeling the cumulative incidence of disease, factors including test kit sensitivity, waning antibody levels, and disease severity can influence seropositivity estimates (1-3). Another important consideration when determining serostatus is the signal-to-cut off (S/Co) threshold used in antibody assays to define seropositive cases. For many commonly used platforms, manufacturer recommended S/Co thresholds were established by testing hospitalized COVID-19 patients, or those with severe illness. Although these thresholds have performed well in populations of similar disease severity, their performance when applied to the general population of those infected with SARS-CoV-2 remains unclear.

We conducted serologic testing to assess the prevalence of SARS-CoV-2-specific antibodies in a cohort of firefighters with a known infection history. We compared the performance sensitivity and specificity of manufacturer recommended S/Co values with modified S/Co values as determined by testing known pre-pandemic negative control samples. We hypothesized that alternate S/Co thresholds would improve performance characteristics and more accurately estimate cumulative incidence of disease in this cohort of firefighters with mild sickness representative of the majority of SARS-CoV-2 infected individuals.

## Methods

The Los Angeles County Department of Public Health Institutional Review Board approved this study. We obtained written informed consent from all study participants. We collected data on participant demographics and polymerase chain reaction (PCR)-confirmed SARS-CoV-2 infection history through electronic surveys.

We collected EDTA plasma venipuncture samples from participants who did not report symptoms on the sample collection day. Serology testing was conducted at Cedars-Sinai Medical Center’s CLIA-certified laboratory with FDA EUA approvals, using the Abbott Architect SARS-CoV-2 assays for IgM and IgG antibodies against spike and nucleocapsid proteins (Abbott Laboratories, Chicago, IL). We classified participants as being seropositive based on two different thresholds: the manufacturer’s recommended S/Co threshold (≥1.4 for IgG; ≥1.0 for IgM) and lower S/Co thresholds based on a validation study that measured antibody levels in 178 negative control patients who had blood drawn prior to the emergence of COVID-19. We summarized descriptive statistics for the study and negative control cohort. We calculated the sensitivity and specificity for manufacturer and modified S/Co thresholds and used percent correctly classified to identify the optimal S/Co threshold based on these samples.

## Results

Overall, 585 firefighters that received an antibody test and had a previous PCR test for SARS-CoV-2. Of these, 52 (8.9%) reported having a PCR-positive test history at a median 3.3 months (Interquartile range [IQR], 1.2-4.2 months) prior to antibody testing. Table 1 presents demographic characteristics for individuals with a known history of infection. Most firefighters were male (90.4%) and between 31-59 years of age (88.5%). The most commonly identified racial/ethnic categories in this cohort included White (48.1%) followed by Hispanic (26.9%), Asian (11.5%), Black or African American (7.7%), and Other (5.8%). There were no firefighters that reported being previously hospitalized for the treatment of COVID-19, indicating that this cohort primarily exhibited mild illness following SARS-CoV-2 infection. For the negative control sample, we measured SARS-CoV-2 antibodies in blood specimens drawn from 178 individuals prior to the emergence of COVID-19. Most individuals in the negative control sample were male (56.2%), white (71.3%), and over 60 years of age (70.2%). The most commonly associated comorbidities described in this cohort included solid tumor cancer (69.1%), hypertension (57.9%), metastatic cancer (32.0%), hypothyroidism (30.3%), and cardiac arrhythmias (28.1%).

**Table 1.** Characteristics of firefighter study participants and negative control cohort.

|  | PCR-Positive<br>Cohort<br>(n=52) | % | Negative<br>Control<br>Sample<br>(n=178) | % | P-value |
| --- | --- | --- | --- | --- | --- |
| Sex |  |  |  |  |  |
| Male | 47 | 90.4% | 100 | 56.2 | <0.001 |
| Female | 5 | 9.6% | 78 | 43.8 |  |
| Age, y |  |  |  |  |  |
| <30 | 5 | 9.6% | 0 | 0.0% | <0.001 |
| 31-44 | 25 | 48.1% | 16 | 9.0% |  |
| 45-59 | 21 | 40.4% | 33 | 18.5% |  |
| ≥60 | 1 | 1.9% | 125 | 70.2% |  |
| Race/ethnicity |  |  |  |  |  |
| Hispanic | 14 | 26.9% | 14 | 7.9% | 0.003 |
| White | 25 | 48.1% | 127 | 71.3% |  |
| Black | 4 | 7.7% | 19 | 10.7% |  |
| Asian | 6 | 11.5% | 19 | 10.7% |  |
| Other | 3 | 5.8% | 10 | 5.6% |  |
| Previous hospitalization for COVID-19 | 0 | 0% | 0 | 0% |  |
| Months between PCR and antibody tests, median (IQR) | 3.3 | 1.2-4.2 | - | - |  |
| Seropositivity (manufacturer threshold ≥ 1.4) | 37 | 71.2% | - | - |  |
| IgM only | 2 | 3.8% | - | - |  |
| IgG only | 12 | 23.1% | - | - |  |
| IgM + IgG | 23 | 44.2% | - | - |  |
| Seropositivity (modified S/Co threshold ≥ 0.36) | 47 | 90.4% | - | - |  |
| IgM only | 1 | 1.9% | - | - |  |
| IgG only | 14 | 26.9% | - | - |  |
| IgM + IgG | 32 | 61.5% | - | - |  |

PCR, polymerase chain reaction; IQR, interquartile range

Thirty-seven (71.2%) firefighters with a previous PCR-positive test were found to be seropositive for SARS-CoV-2 IgG and/or IgM antibodies based on the manufacturer recommended S/Co thresholds. This corresponded to a sensitivity of 71.2% (95% confidence interval [CI], 56.9-82.9%) and specificity of 100%. We then tested the performance of several modified S/Co thresholds (1.0, 0.60, 0.36, 0.34, 0.30, 0.26, 0.18), based on the index titer values measured in the negative control sample (Figure 1). As expected, we observed a general improvement in test sensitivity with lowering S/Co thresholds at the expense of declining specificity (Table 2). We found that a modified S/Co of 0.36 yields optimal sensitivity and specificity performance (percent correctly classified=97.0%) in this cohort (Table 2). When applying this modified threshold to estimate overall serostatus in firefighters, 47 (90.4%) individuals with a previous PCR-positive test result were determined to be seropositive.

**Table 2.** Analysis to determine optimal S/Co threshold.

| S/Co threshold | Specificity (based on negative sample) | Sensitivity (based on PCR+ sample) | Percent correctly classified |
| --- | --- | --- | --- |
| 1.4 | 100% | 71.2% (56.9-82.9%) | 92.6% |
| 1.0 | 99.4% (96.9-99.9%) | 75.0% (61.1-86.0%) | 93.5% |
| 0.60 | 99.4% (96.9-99.9%) | 76.9% (63.2-87.5%) | 93.9% |
| 0.36 | 99.4% (96.9-99.9%) | 90.4% (79.0-96.8%) | 97.0% |
| 0.34 | 98.3% (95.2-99.7%) | 90.4% (79.0-96.8%) | 96.1% |
| 0.30 | 97.8% (94.4-99.4%) | 90.4% (79.0-96.8%) | 95.7% |
| 0.26 | 97.2% (93.6-99.1%) | 90.4% (79.0-96.8%) | 95.2% |
| 0.18 | 96.1% (92.1-98.4%) | 94.2% (84.1-98.8%) | 95.2% |

**Figure 1.**
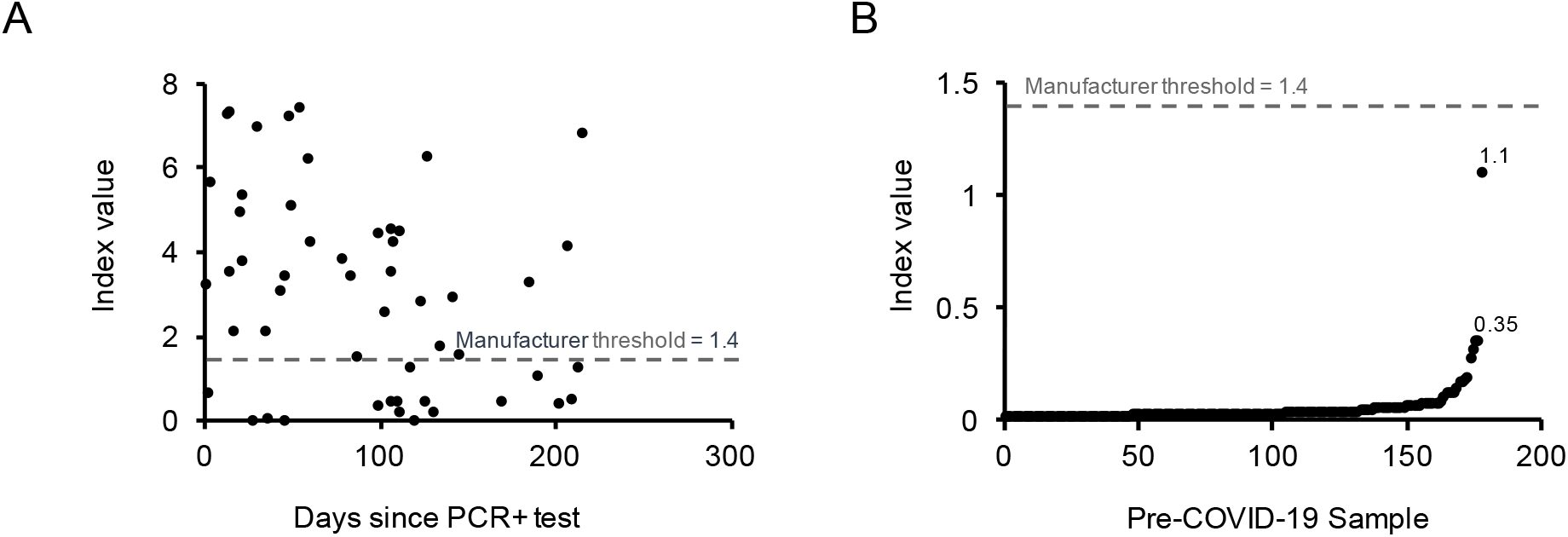
Index values of SARS-CoV-2 antibody titers measured in (A) serum collected from firefighters shown by days since PCR test positivity and (B) negative control serum collected from hospitalized individuals prior to COVID-19. The gray line denotes the manufacturer recommended S/Co cut off.

## Discussion

Serologic tests can be used to identify active and resolved SARS-CoV-2 infections, which is critical for the epidemiological tracking of COVID-19 in the population. Manufacturer recommended S/Co thresholds for serologic tests are typically based on samples collected from symptomatic patients within a few weeks of infection. While these cut-offs have been shown to perform well in cohorts of similar disease severity, it has been suggested that these thresholds may underestimate cumulative incidence when applied to individuals with milder disease severity or as antibodies wane overtime (2, 3). Previous analysis of the same Abbott Architect instrument utilized in this study shows that manufacturer based S/Co thresholds were highly sensitive when applied to samples collected from hospitalized patients assayed within 17 days of PCR positivity (4). However, owing to the highly selective characteristics of the sensitivity cohort evaluated in their study, the authors speculate that modified S/Co thresholds may be considered for diagnostic serology in different target populations (4).

To this end, we evaluated the performance of the Abbott Architect using manufacturer recommended S/Co thresholds in a cohort of firefighters with a known history of SARS-CoV-2 infection. In contrast to previous studies evaluating the performance characteristics of this instrument, none of the firefighters included in the present analysis were hospitalized for the treatment of COVID-19. Our data demonstrate that the sensitivity of manufacturer S/Co thresholds significantly underperforms in ambulatory patients, particularly with increasing time since initial PCR positive test. As a result, we pursued alternate S/Co thresholds based on IgG-antibody titer index values measured in known negative samples collected from pre-COVID-19 serum. Analysis of the percent correctly classified according to multiple alternate S/Co thresholds determined the optimal S/Co threshold to be 0.36. The sensitivity and specificity of this threshold was 90.4% (95% CI, 79.0-96.8%) and 99.4% (95% CI, 96.9-99.9%), respectively. When applied to the firefighter cohort, this threshold improved the overall seroprevalence estimate by 19.2% compared to the manufacturer recommended S/Co threshold. Given that approximately 85% of individuals infected by SARS-CoV-2 are not expected to be hospitalized for the management of their disease, our data suggest that lowering S/Co thresholds from manufacturer recommended levels may be more optimal for antibody assessments in the general population (5).

Overall, our data demonstrate that the use of a modified S/Co based on known negative samples significantly increases the percent seropositive and more accurately estimates cumulative incidence of infection in a cohort with a disease severity largely representative of the general COVID-19 population. These results are consistent with evidence from studies from US and Belgium which find detectable antibodies several months after infection (6, 7). In summary, our findings suggest that estimation of cumulative incidence of COVID-19 using serology-based assays must apply diagnostic thresholds that account for weaker antibody response exhibited by those with mild disease.

The findings of this study should be viewed in light of its limitations. First, seroprevalence was assessed based on blood specimens drawn at a single, cross-sectional time point, resulting in varying times since initial PCR positivity. Future serology studies evaluating S/Co thresholds may benefit from repeated measurements to longitudinally track antibody kinetics over time. In addition, PCR positivity was determined by participant survey, thus we cannot rule out false-positive PCR test histories. Finally, it is unclear whether factors such as age, race/ethnicity, concomitant comorbidities, and cross-reactivity with other known coronaviruses influenced SARS-CoV-2 antibody measurements in the negative control cohort utilized in this study. Additional studies evaluating SARS-CoV-2 antibody levels in larger and more representative negative control specimens are needed.

## Data Availability

The primary data included in the manuscript may be available upon request through email of the corresponding author of the research team.

## References

1. Takahashi S, Greenhouse B, Rodríguez-Barraquer I. Are Seroprevalence Estimates for Severe Acute Respiratory Syndrome Coronavirus 2 Biased? The Journal of Infectious Diseases. 2020;222(11):1772–5.

2. Ibarrondo FJ, Fulcher JA, Goodman-Meza D, Elliott J, Hofmann C, Hausner MA, et al. Rapid Decay of Anti–SARS-CoV-2 Antibodies in Persons with Mild Covid-19. New England Journal of Medicine. 2020;383(11):1085–7.

3. Seow J, Graham C, Merrick B, Acors S, Pickering S, Steel KJA, et al. Longitudinal observation and decline of neutralizing antibody responses in the three months following SARS-CoV-2 infection in humans. Nature Microbiology. 2020 2020/12/01;5(12):1598–607.

4. Bryan A, Pepper G, Wener MH, et al. Performance characteristics of the Abbott Architect SARS-CoV-2 IgG assay and seroprevalence in Boise, Idaho. J. Clin. Microbiol. 2020;58(8):e00941–20.

5. Nakamichi K, Shen JZ, Lee CS, et al. Hospitalization and mortality associated with SARS-CoV-2 viral clades in COVID-19. Sci Rep. 2021;11(1):4802.

6. Duysburgh E, Mortgat L, Barbezange C, Dierick K, Fischer N, Heyndrickx L, et al. Persistence of IgG response to SARS-CoV-2. The Lancet Infectious Diseases. 2021;21(2):163–4.

7. Wajnberg A, Amanat F, Firpo A, Altman DR, Bailey MJ, Mansour M, et al. Robust neutralizing antibodies to SARS-CoV-2 infection persist for months. Science. 2020;370(6521):1227–30.

